# HEalth literacy in Low back Pain – the HELP! media intervention study: A stepped wedge cluster randomised trial

**DOI:** 10.64898/2025.12.11.25342096

**Authors:** Mathangi Shanthakumar, Laurent Billot, Manuela Ferreira, HELP study Investigators

## Abstract

The HELP study aims to evaluate the effectiveness of a media-led intervention that includes evidence-based information on the management of low back pain (LBP), distributed and broadcasted at point-of-care in primary care settings (i.e. General Practice clinic waiting rooms), to change beliefs and behaviour about LBP management.

The effect of the intervention on patients’ beliefs and attitudes about LBP management will be evaluated using The Back Beliefs Questionnaire (BBQ) and the Back Pain Attitudes Questionnaire (Back-PAQ) scores. Change in medical practitioner behaviour and delivery of care (i.e. prescription of opioid medication, referrals for medical imaging and allied health care) will be assessed using administrative data.

The statistical analysis plan pre-specifies the method of analysis for the key outcome variables collected in the study. The BBQ and Back-PAQ scores will estimate the mean difference (MD) in each score between the intervention arm and the usual care arm obtained from a linear mixed model. Using a similar model, analysis of the GP clinic data will report the differences in proportions of opioid medication, and referrals to medical imaging and allied health care between the intervention arm and usual care arm.

## 2 Introduction

### 2.1 Study synopsis

The HELP study is a multicentre, stepped-wedge, cluster randomised trial with the key objectives to:

- Evaluate the effectiveness of a media-led intervention that includes evidence-based information on the management of low back pain (LBP), distributed and broadcasted at point-of-care in primary care settings (i.e. General Practice clinic waiting rooms), to change patient beliefs and attitudes about LBP.
- Determine the effectiveness of the media-led intervention in changing Medical Practitioner behaviour and practice in delivery of care (i.e. medication prescription, referrals for medical imaging and specialist or allied health care).

Thirty large participating General Practice (GP) clinics from 4 Primary Health Networks (PHNs) were randomised into 5 sequences to receive the intervention, using a stepped-wedge design with 6 periods (5 steps). All practices commenced the study simultaneously on 17 June 2024 and progressed simultaneously through six 4-week periods ending on 1 December 2024, thus resulting in a total of 6-month duration. During the first period, all GPs were under usual care (control). At each subsequent period, one group of GPs crossed from control to intervention until all groups switched to the intervention by Period 6.

**Figure 1.**
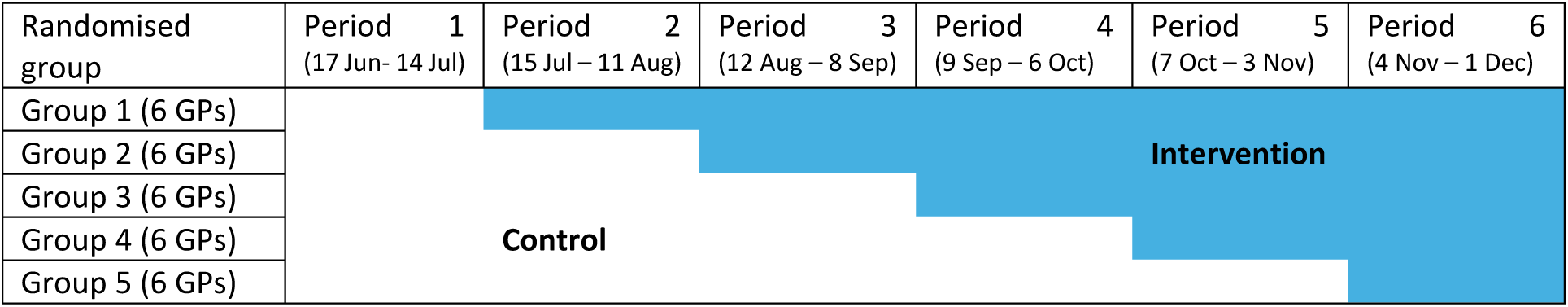
Study schema

### 2.2 Study population

#### 2.2.1 Inclusion and exclusion criteria

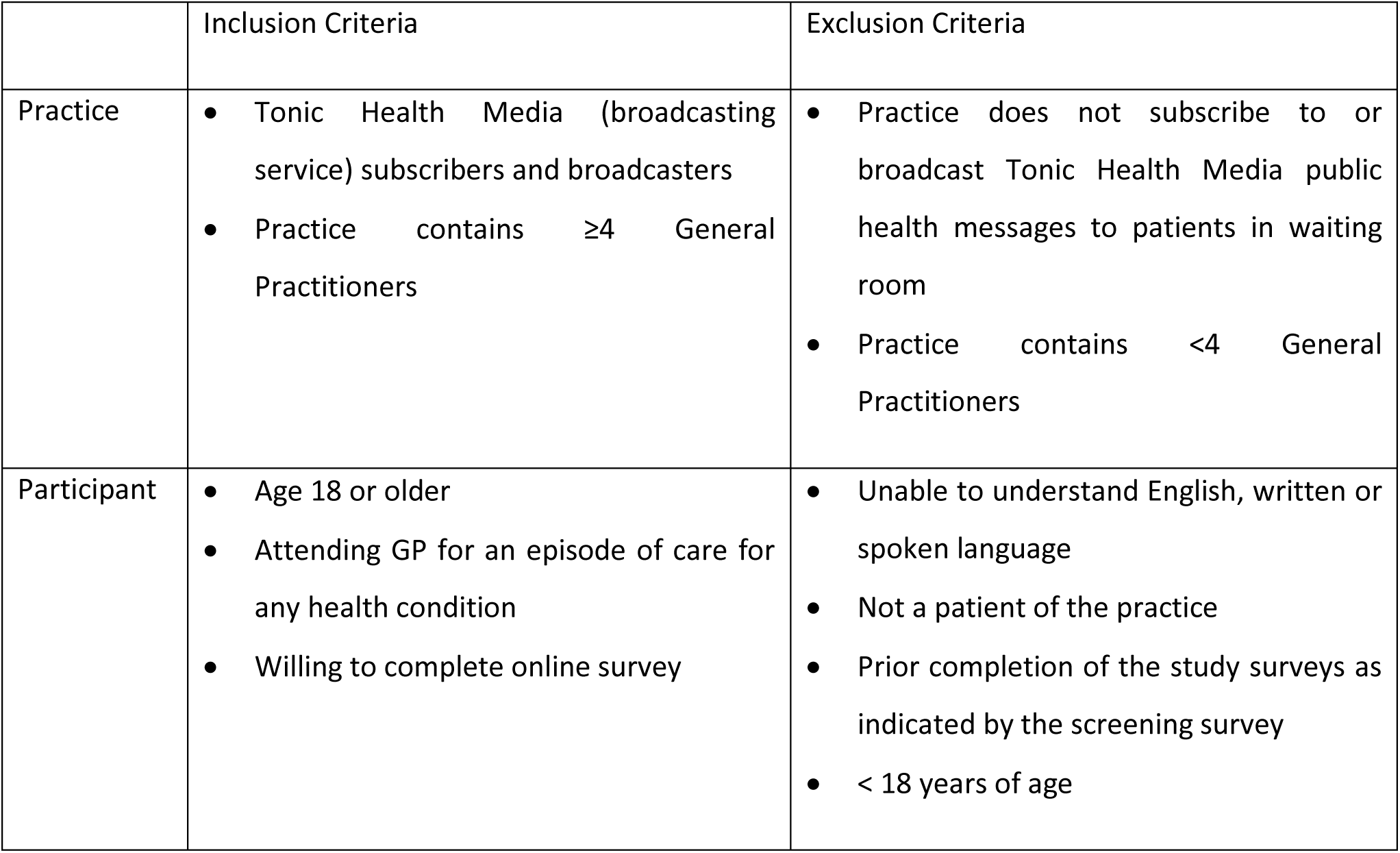

2.2 Study population 2.2.1 Inclusion and exclusion criteria
|  | Inclusion Criteria | Exclusion Criteria |
| --- | --- | --- |
| Practice | • Tonic Health Media (broadcasting service) subscribers and broadcasters<br>• Practice contains $\geq 4$ General Practitioners | • Practice does not subscribe to or broadcast Tonic Health Media public health messages to patients in waiting room<br>• Practice contains $< 4$ General Practitioners |
| Participant | • Age 18 or older<br>• Attending GP for an episode of care for any health condition<br>• Willing to complete online survey | • Unable to understand English, written or spoken language<br>• Not a patient of the practice<br>• Prior completion of the study surveys as indicated by the screening survey<br>• $< 18$ years of age |

### 2.3 Intervention and Control

During the study periods, GP waiting room screens will display the study digital advertising poster and educational videos.

- **Intervention periods** will display educational videos on LBP with six key messages from international clinical practice guidelines.
- **Control periods** will display educational videos on other health conditions, but no information on LBP management will be offered.

The study posters and pamphlets will contain a brief overview of the study and a QR code for participants to access the online participant information sheet, pre-screening form, and consent form. Data about baseline characteristics, their beliefs and attitudes about LBP, and feedback will be collected from the participants.

The days and times the intervention video is displayed at the participating practices will be recorded and participants in the intervention period asked to confirm whether they have watched the video while waiting prior to completing the survey online.

### 2.4 Outcome measures

#### 2.4.1 Participant-level outcome measures

Outcome measures of beliefs and attitudes about LBP were collected from the questionnaire completed by the participant during their one^1^ GP visit during either the intervention or control period.

- **Beliefs about back pain will be measured using** the 14 item Back Beliefs Questionnaire (BBQ) [1] – total scores range between 9 and 45, with higher scores indicating more positive beliefs and better ability to cope.
- **Attitudes about back pain will be measured using** the 20-item Back Pain Attitudes Questionnaire (Back-PAQ) [2] – total scores range between 20 and 100, with higher scores indicating more unhelpful beliefs.

#### 2.4.2 Practice-level outcome measures

De-identified data of those who were reviewed for LBP from the participating GP clinics during the study period will be used to prepare practice-level outcome measures.

Prior to unblinding, the practice-level outcome measures from the full list specified in the Protocol were narrowed down to three key meaningful ones of interest. All other practice-level outcome measures will be reported descriptively, with no formal tests applied to them.

The three key practice-level outcome measures are:

- Prescription of opioid medication
- Referral for any medical imaging (i.e. X-ray, CT scan, MRI, Bone scan, Other)
- Referral to any allied health care (i.e. Physiotherapy, Occupational Therapy, Exercise Physiology, Dietetics and Chiropractic)

The other practice-level outcome measures are:

- Each type of medical prescriptions for LBP, other than opioid (i.e. anti-inflammatory, pain relief, pregabalin, anti-depressants, sleeping pill, and other prescriptions)
- Each type of referral for medical imaging (i.e. X-ray, CT scan, MRI, Bone scan, Other)
- Each type of referral to specialist review (i.e. Rheumatologist, Orthopaedic surgeon, Neurosurgeon, Pain specialist, Other)
- Each type of referral to allied health care (i.e. Physiotherapy, Occupational Therapy, Exercise Physiology, Dietetics and Chiropractic)

GP referrals and prescriptions for LBP will be collected over a 12-month period, being:

- Pre – 3 months before the intervention period (25 March 2024 – 16 June 2024)
- Mid – during the intervention period (17 June 2024 – 1 December 2024)
- Post – 3 months after the intervention period (2 December 2024 – 23 February 2025)

Those in the ‘Pre’-period will be included in the Control group for analyses, and those in the ‘Post’-period will be included in the Intervention group for analyses.

### 2.5 Randomisation and blinding

**Randomisation** of the 30 participating GP practices to one of 5 possible sequences was done by an independent researcher using a concealed list of anonymised codes for each GP and sequence. This researcher also determined the order of sequences to receive the intervention. Participants are not randomised, and those who have already participated will be ineligible to participate a second time.

**Blinding** the allocation from GPs and clinic staff will not be possible due to the nature of the intervention, however researchers as well as the planning and initial programming of analyses will be done blinded to the random allocation.

### 2.6 Statistical hypothesis

The statistical hypotheses for participant-level survey data are as follows:

- **Null hypothesis 1**: no difference in the Back Beliefs Questionnaire (BBQ) scores between those randomised to Intervention and Control, that is mean difference (MD) (Intervention vs Control) = 0
- **Null Hypothesis 2**: no difference in the Back Pain Attitudes Questionnaire (Back-PAQ) scores between those randomised to Intervention and Control, that is mean difference (MD) (Intervention vs Control) = 0
- **Alternative hypothesis (2-sided)**: MD (Intervention vs Control) ≠ 0, with all previous hypotheses rejected below their respective thresholds.

The statistical hypotheses for practice-level GP data are as follows:

- **Null hypothesis 1**: no difference in the GP prescription of opioid medication between those randomised to Intervention and Control, that is odds ratio (OR) (Intervention vs Control) = 1
- **Null hypothesis 2**: no difference in the GP patients referred for any medical imaging between those randomised to Intervention and Control, that is odds ratio (OR) (Intervention vs Control) = 1
- **Null hypothesis 3**: no difference in the GP patients referred to any allied health care between those randomised to Intervention and Control, that is odds ratio (OR) (Intervention vs Control) = 1
- **Alternative hypothesis (2-sided)**: OR (Intervention vs Control) ≠ 0, with all previous hypotheses rejected below their respective thresholds.

### 2.7 Sample size

The study was designed as a stepped-wedge trial design with a cross-sectional sampling structure and an exchangeable correlation structure, not allowing for varying cluster sizes, with 5 sequences/steps and 6 clusters per sequence, assuming:

– an intra-cluster correlation (ICC) of 0.03,
– for a continuous outcome with a mean difference of 0.2 standard deviations (equivalent to Cohen’s d = 0.2),
– a normal approximation with significance level of 0.05,
– and a desired power of 90%,

The required cluster size per period was initially calculated as 5 participants, leading to a target total sample size of about 900 participants (5 randomised sequences, each containing 6 GPs with each GP recruiting 5 participants in each of the 6 periods i.e. 5×6×5×6 = 900).

However, upon review of the calculation, we realised there was a mistake in the original sample size calculation and that to achieve 90% power, each practice had to recruit 16 participants per period, not 5 as initially anticipated, thus requiring a total sample size of about 2,880 participants (5 sequences x 6 GPs x 6 periods x 16 participants = 2,880 participants).

We will perform all analyses as intended using models for the analysis of stepped-wedge trials while acknowledging the reduced statistical power.

## 3 Statistical analysis

### 3.1 Statistical principles

#### 3.1.1 Level of statistical significance

Final analyses will be conducted using a simple step-down approach, the Holm-Bonferroni method, separately for participant-level and practice-level outcomes to control for the family-wise error rate (FWER) when performing multiple comparisons.

#### 3.1.2 Statistical software

Analyses will be conducted primarily using SAS Enterprise Guide version 8.3 or R version 4.5.1.

### 3.2 Analysis populations

Due to the stepped-wedge design, the group allocation for a patient is determined by the practice and by the period during which they participated, regardless of treatment adherence.

The intention-to-treat analysis set will be used to assess both effectiveness and safety. The flow of patients through the study will be displayed in a CONSORT diagram (Figure 1). Analyses will be undertaken according to reporting guidelines for stepped-wedge cluster randomised trials [3].

### 3.3 Baseline analyses

#### 3.3.1 Cluster characteristics

Description of the cluster characteristics (e.g. location, size) will be presented by treatment group. Discrete variables will be summarised by frequencies and percentages. Percentages will be calculated according to the number of clusters with available data. Continuous variables will be summarised by using mean and standard deviation (SD), and median and interquartile range (Q1-Q3).

#### 3.3.2 Patient characteristics

Description of the baseline characteristics will be presented by treatment group and by study period. Discrete variables will be summarised by frequencies and percentages. Percentages will be calculated according to the number of patients in whom data are available. Continuous variables will be summarised by using mean and SD, and median and interquartile range (Q1-Q3). No adjustment for clustering will be applied when summarising baseline characteristics. Baseline measures will include all socio-demographic, beliefs, attitudes and feedback collected at baseline.

### 3.4 Analysis of the participant-level outcomes

The participant-level outcome measures of the BBQ score (of 9 to 45) and the Back-PAQ score (of 20 to 100) will be analysed independently as continuous variables. The intervention effect for each of the two outcomes will be estimated as the mean difference (MD) between the intervention arm and the usual care arm obtained from a linear mixed model, assuming a normal distribution and an identity link function (defined below).

A simple sequentially rejective multiple test procedure will be used [4], whereby basic hypotheses are rejected one at a time according to certain rules, until no further rejections can be done.

For the two participant-level outcomes (i.e. the BBQ and Back-PAQ scores):

– If the smaller of the two p-values is less than 0.025 (α/2) then it is considered significant. We reject the first hypothesis, and we can test the larger of the two p-values at the 5% (α) level.
– If the larger of the two p-values is less than 0.05 then it is also considered significant.

At any point when the p-value is greater than the threshold, we stop.

#### 3.4.1 Analysis model

The analysis will be conducted using the approach developed by Hussey and Hughes for the analysis of stepped-wedge trials [5]. We will be using a linear mixed-effects model with a random effect for cluster (GP clinic), and a fixed effect indicating the group assignment of each cluster at each period, and a fixed categorical effect of time (each period). The corresponding linear mixed model can be written as follows:

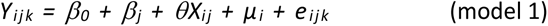

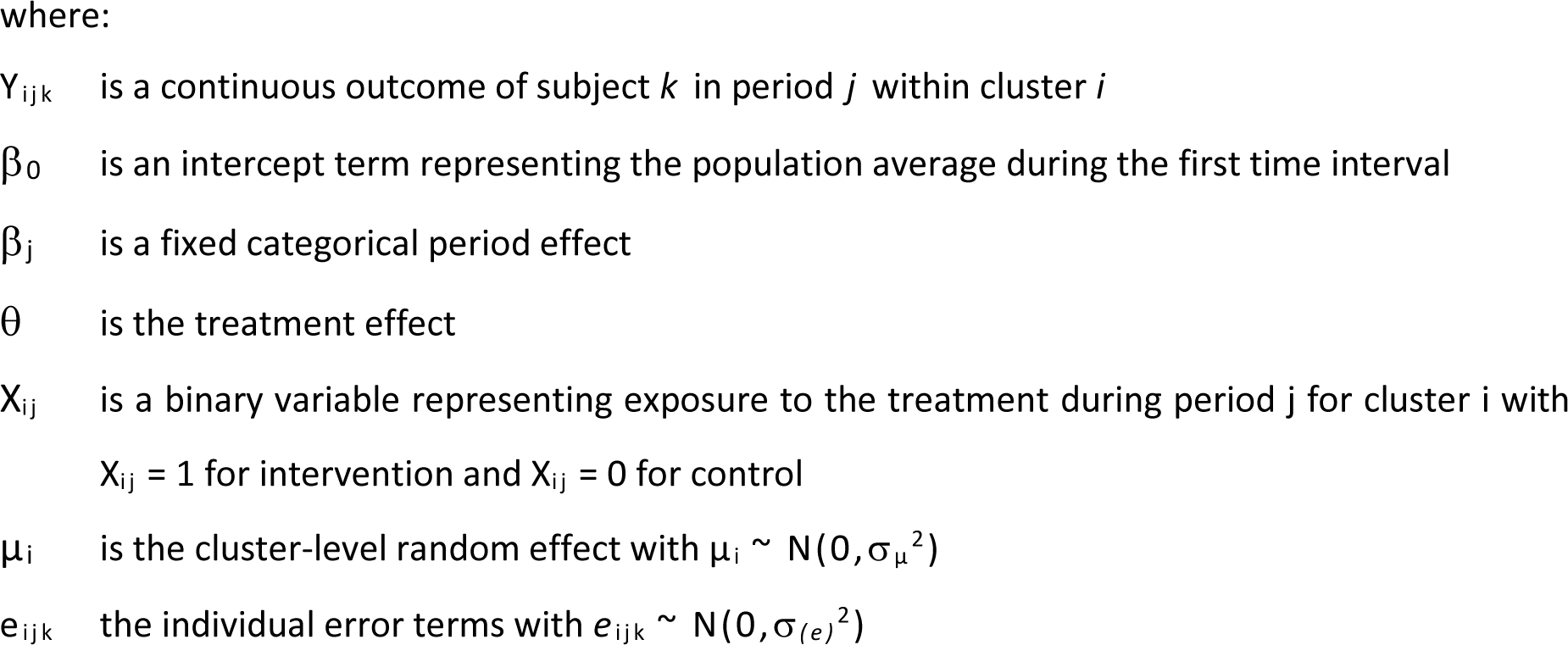

For the BBQ score, the effect of the intervention will be presented as the MD of a better outcome and its 95% confidence interval (CI) using the control arm as the reference (i.e. where a positive MD corresponds to a better BBQ score in the intervention arm compared to the control arm).

For the Back-PAQ score, the effect of the intervention will be presented as the MD of a worse outcome and its 95% confidence interval (CI) using the control arm as the reference (i.e. where a positive MD corresponds to a worse Back-PAQ score in the intervention arm compared to the control arm).

#### 3.4.2 Adjusted analyses

The model described in Section 3.4.1 will be re-run after adding the following individual covariates as fixed effects:

- Age (continuous)
- Sex (male vs female)
- Presently have low back pain

#### 3.4.3 Treatment of missing data

The proportion of data missing for the outcomes (BBQ score and Back-PAQ) will be described overall, by study period. In case of non-negligible amounts of missing data (>5%) or changes in missing data patterns overtime, we will use controlled multiple imputations to assess under what conditions the results change, and how plausible these conditions are, using the approach described by Cro et al. [6]

### 3.5 Analysis of the practice-level outcomes

GP extracted information is de-identified and provided in either patient-level or aggregated format from the data custodians. All patient-level data will be aggregated in 4-week periods over the 3 time points: 3 months pre, mid, and 3 months post the delivery of the study intervention, in line with the requested format of the aggregated data.

The proportions of opioid medication prescriptions, referrals for any medical imaging, and referrals to any allied health care will compare the intervention and control groups using a generalised linear mixed effect model with Poisson link function (using a similar approach to 3.4.1). Over-dispersion in the Poisson model will be tested using Wald’s test or Likelihood Ratio Test (LRT), with a negative binomial link function to be used in the generalised mixed model if the formal test indicates over-dispersion. Statistical significance will be defined as p<0.05 on the basis of a two-sided test.

A simple sequentially rejective multiple test procedure will be used, whereby basic hypotheses are rejected one at a time according to certain rules, until no further rejections can be done.

For the three key practice-level outcomes (i.e. the number of opioid prescriptions, referrals for any medical imaging, and referrals to any allied health care):

– If the smallest of the three p-values is less than 0.0167 (α/3) then it is considered significant. We reject the first hypothesis, and we can test the next smallest of the p-values at the 2.5% (α/2) level.
– If the second smallest p-value is less than 0.025 then it is considered significant. We reject the second hypothesis, and we can test the largest of the three p-values at the 5% (α) level.
– If the largest of the three p-values is less than 0.05 then it is also considered significant.

At any point when the p-value is greater than the threshold, we stop.

## Data Availability

All participant data collected by the study will be de-identified and analysed according to group i.e. intervention or control arm.
GP data will be de-identified for all patients attending for a LBP review and will report the total number of prescriptions of opioid medication, total number of referrals for medical imaging, and total number of referrals to allied health care.
No individual participant data will be publicly available to protect confidentially and privacy of individual participants.

## 5. Proposed outputs

### 5.1 Tables

**Table 1.** Number of subjects enrolled and with participant-level outcome data per group and per period.

| Group | Period 0<br>17/06/24-14/07/24 | Period 1<br>15/07/24-11/08/24 | Period 2<br>12/08/24-08/09/24 | Period 3<br>09/09/24-06/10/24 | Period 4<br>07/10/24-03/11/24 | Period 5<br>04/11/24-01/12/24 | Total |
| --- | --- | --- | --- | --- | --- | --- | --- |
| **1 (xx clusters)** | xxx / xxx (xx%) | xxx / xxx (xx%) | xxx / xxx (xx%) | xxx / xxx (xx%) | xxx / xxx (xx%) | xxx / xxx (xx%) | xxx / xxx (xx%) |
| **2 (xx clusters)** | xxx / xxx (xx%) | xxx / xxx (xx%) | xxx / xxx (xx%) | xxx / xxx (xx%) | xxx / xxx (xx%) | xxx / xxx (xx%) | xxx / xxx (xx%) |
| **3 (xx clusters)** | xxx / xxx (xx%) | xxx / xxx (xx%) | xxx / xxx (xx%) | xxx / xxx (xx%) | xxx / xxx (xx%) | xxx / xxx (xx%) | xxx / xxx (xx%) |
| **4 (xx clusters)** | xxx / xxx (xx%) | xxx / xxx (xx%) | xxx / xxx (xx%) | xxx / xxx (xx%) | xxx / xxx (xx%) | xxx / xxx (xx%) | xxx / xxx (xx%) |
| **5 (xx clusters)** | xxx / xxx (xx%) | xxx / xxx (xx%) | xxx / xxx (xx%) | xxx / xxx (xx%) | xxx / xxx (xx%) | xxx / xxx (xx%) | xxx / xxx (xx%) |
| **Total (xx clusters)** | xxx / xxx (xx%) | xxx / xxx (xx%) | xxx / xxx (xx%) | xxx / xxx (xx%) | xxx / xxx (xx%) | xxx / xxx (xx%) | xxx / xxx (xx%) |

Notes:
The denominator represents the number of subjects enrolled during the period.
The numerator represents the number of subjects with a participant-level outcome measure.
Cells shaded in grey represent control/no intervention.
Cells shaded in blue represent intervention provided.

**Table 2.** Number of subjects reviewed for LBP at a participating GP clinic and with any of the key practice-level outcome data per group and per period.

| Group | Pre-period<br>25/03/24-16/06/24 | Period 0<br>17/06/24-14/07/24 | Period 1<br>15/07/24-11/08/24 | Period 2<br>12/08/24-08/09/24 | Period 3<br>09/09/24-06/10/24 | Period 4<br>07/10/24-03/11/24 | Period 5<br>04/11/24-01/12/24 | Post-period<br>02/12/24-23/02/25 | Total |
| --- | --- | --- | --- | --- | --- | --- | --- | --- | --- |
| **1 (xx clusters)** | xxx / xxx<br>(xx%) | xxx / xxx<br>(xx%) | xxx / xxx<br>(xx%) | xxx / xxx<br>(xx%) | xxx / xxx<br>(xx%) | xxx / xxx<br>(xx%) | xxx / xxx<br>(xx%) | xxx / xxx<br>(xx%) | xxx / xxx<br>(xx%) |
| **2 (xx clusters)** | xxx / xxx<br>(xx%) | xxx / xxx<br>(xx%) | xxx / xxx<br>(xx%) | xxx / xxx<br>(xx%) | xxx / xxx<br>(xx%) | xxx / xxx<br>(xx%) | xxx / xxx<br>(xx%) | xxx / xxx<br>(xx%) | xxx / xxx<br>(xx%) |
| **3 (xx clusters)** | xxx / xxx<br>(xx%) | xxx / xxx<br>(xx%) | xxx / xxx<br>(xx%) | xxx / xxx<br>(xx%) | xxx / xxx<br>(xx%) | xxx / xxx<br>(xx%) | xxx / xxx<br>(xx%) | xxx / xxx<br>(xx%) | xxx / xxx<br>(xx%) |
| **4 (xx clusters)** | xxx / xxx<br>(xx%) | xxx / xxx<br>(xx%) | xxx / xxx<br>(xx%) | xxx / xxx<br>(xx%) | xxx / xxx<br>(xx%) | xxx / xxx<br>(xx%) | xxx / xxx<br>(xx%) | xxx / xxx<br>(xx%) | xxx / xxx<br>(xx%) |
| **5 (xx clusters)** | xxx / xxx<br>(xx%) | xxx / xxx<br>(xx%) | xxx / xxx<br>(xx%) | xxx / xxx<br>(xx%) | xxx / xxx<br>(xx%) | xxx / xxx<br>(xx%) | xxx / xxx<br>(xx%) | xxx / xxx<br>(xx%) | xxx / xxx<br>(xx%) |
| **Total (xx clusters)** | xxx / xxx<br>(xx%) | xxx / xxx<br>(xx%) | xxx / xxx<br>(xx%) | xxx / xxx<br>(xx%) | xxx / xxx<br>(xx%) | xxx / xxx<br>(xx%) | xxx / xxx<br>(xx%) | xxx / xxx<br>(xx%) | xxx / xxx<br>(xx%) |

**Notes:**
The denominator represents the number of subjects reviewed for LBP during the period.
The numerator represents the number of subjects with any of the key practice-level outcome measure.
Cells shaded in grey represent control/no intervention.
Cells shaded in blue represent intervention provided.

**Table 3.** Baseline characteristics.

|  | Intervention<br>(N = ) | Control<br>(N = ) | Total<br>(N = ) |
| --- | --- | --- | --- |
| **Age (years)** |  |  |  |
| N | xxx | xxx | xxx |
| Mean (SD) | xx.x (xx.x) | xx.x (xx.x) | xx.x (xx.x) |
| Median (Q1; Q3) | xx.x (xx.x; xx.x) | xxx (xxx; xxx) | xxx (xxx; xxx) |
| Min Max | xxx xxx | xxx xxx | xxx xxx |
| **Sex** |  |  |  |
| Male | xxx/xxx (xx.x%) | xxx/xxx (xx.x%) | xxx/xxx (xx.x%) |
| Female | xxx/xxx (xx.x%) | xxx/xxx (xx.x%) | xxx/xxx (xx.x%) |
| **Reason for GP visit** |  |  |  |
| I have an appointment to discuss my health | xxx/xxx (xx.x%) | xxx/xxx (xx.x%) | xxx/xxx (xx.x%) |
| I am accompanying a relative or friend | xxx/xxx (xx.x%) | xxx/xxx (xx.x%) | xxx/xxx (xx.x%) |
| I have come to book an appointment | xxx/xxx (xx.x%) | xxx/xxx (xx.x%) | xxx/xxx (xx.x%) |
| I have come for some health-related tests | xxx/xxx (xx.x%) | xxx/xxx (xx.x%) | xxx/xxx (xx.x%) |
| Other | xxx/xxx (xx.x%) | xxx/xxx (xx.x%) | xxx/xxx (xx.x%) |
| **Previously visited a GP for this condition** |  |  |  |
| Yes, and within the last 48 hours | xxx/xxx (xx.x%) | xxx/xxx (xx.x%) | xxx/xxx (xx.x%) |
| Yes, but not within the last 48 hours | xxx/xxx (xx.x%) | xxx/xxx (xx.x%) | xxx/xxx (xx.x%) |
| No | xxx/xxx (xx.x%) | xxx/xxx (xx.x%) | xxx/xxx (xx.x%) |
| **Presently have low back pain** |  |  |  |
| Yes | xxx/xxx (xx.x%) | xxx/xxx (xx.x%) | xxx/xxx (xx.x%) |
| No | xxx/xxx (xx.x%) | xxx/xxx (xx.x%) | xxx/xxx (xx.x%) |
| **Highest level of education** |  |  |  |
| Year 10 or below | xxx/xxx (xx.x%) | xxx/xxx (xx.x%) | xxx/xxx (xx.x%) |
| Year 12 | xxx/xxx (xx.x%) | xxx/xxx (xx.x%) | xxx/xxx (xx.x%) |
| TAFE or apprenticeship | xxx/xxx (xx.x%) | xxx/xxx (xx.x%) | xxx/xxx (xx.x%) |
| Graduate certificate | xxx/xxx (xx.x%) | xxx/xxx (xx.x%) | xxx/xxx (xx.x%) |
| Degree | xxx/xxx (xx.x%) | xxx/xxx (xx.x%) | xxx/xxx (xx.x%) |
| Masters | xxx/xxx (xx.x%) | xxx/xxx (xx.x%) | xxx/xxx (xx.x%) |
| PhD | xxx/xxx (xx.x%) | xxx/xxx (xx.x%) | xxx/xxx (xx.x%) |
| Other | xxx/xxx (xx.x%) | xxx/xxx (xx.x%) | xxx/xxx (xx.x%) |
| **Relationship status** |  |  |  |
| Single | xxx/xxx (xx.x%) | xxx/xxx (xx.x%) | xxx/xxx (xx.x%) |
| Married | xxx/xxx (xx.x%) | xxx/xxx (xx.x%) | xxx/xxx (xx.x%) |
| De Facto | xxx/xxx (xx.x%) | xxx/xxx (xx.x%) | xxx/xxx (xx.x%) |
| Rather not say | xxx/xxx (xx.x%) | xxx/xxx (xx.x%) | xxx/xxx (xx.x%) |
| Other | xxx/xxx (xx.x%) | xxx/xxx (xx.x%) | xxx/xxx (xx.x%) |
| **Work status** |  |  |  |
| Full time carer (including parent) | xxx/xxx (xx.x%) | xxx/xxx (xx.x%) | xxx/xxx (xx.x%) |
| Part time work | xxx/xxx (xx.x%) | xxx/xxx (xx.x%) | xxx/xxx (xx.x%) |
| Full time work | xxx/xxx (xx.x%) | xxx/xxx (xx.x%) | xxx/xxx (xx.x%) |
| Volunteer | xxx/xxx (xx.x%) | xxx/xxx (xx.x%) | xxx/xxx (xx.x%) |
| Retired | xxx/xxx (xx.x%) | xxx/xxx (xx.x%) | xxx/xxx (xx.x%) |
| Unemployed | xxx/xxx (xx.x%) | xxx/xxx (xx.x%) | xxx/xxx (xx.x%) |
| Studying | xxx/xxx (xx.x%) | xxx/xxx (xx.x%) | xxx/xxx (xx.x%) |
| Other | xxx/xxx (xx.x%) | xxx/xxx (xx.x%) | xxx/xxx (xx.x%) |

Notes:

**Table 4.**
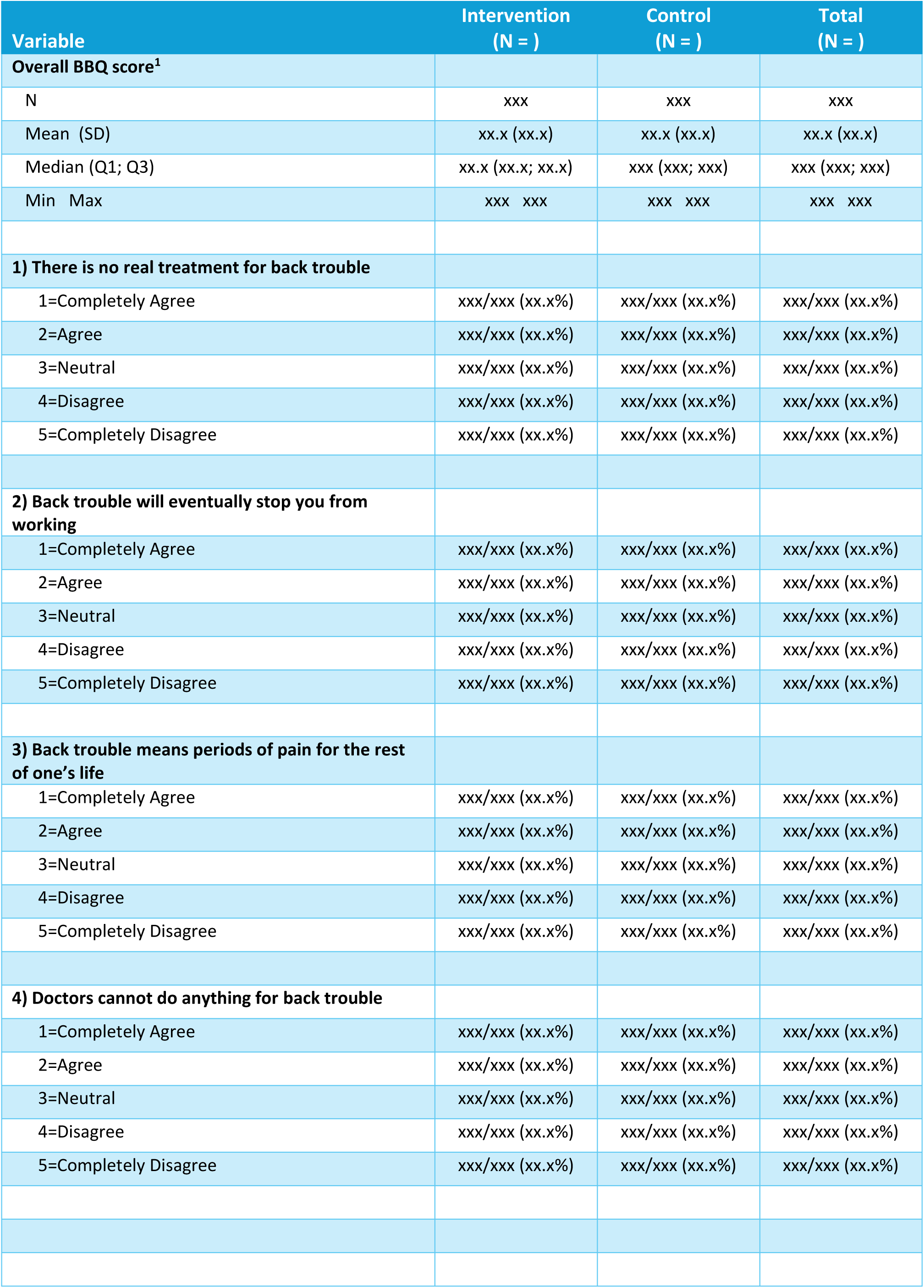
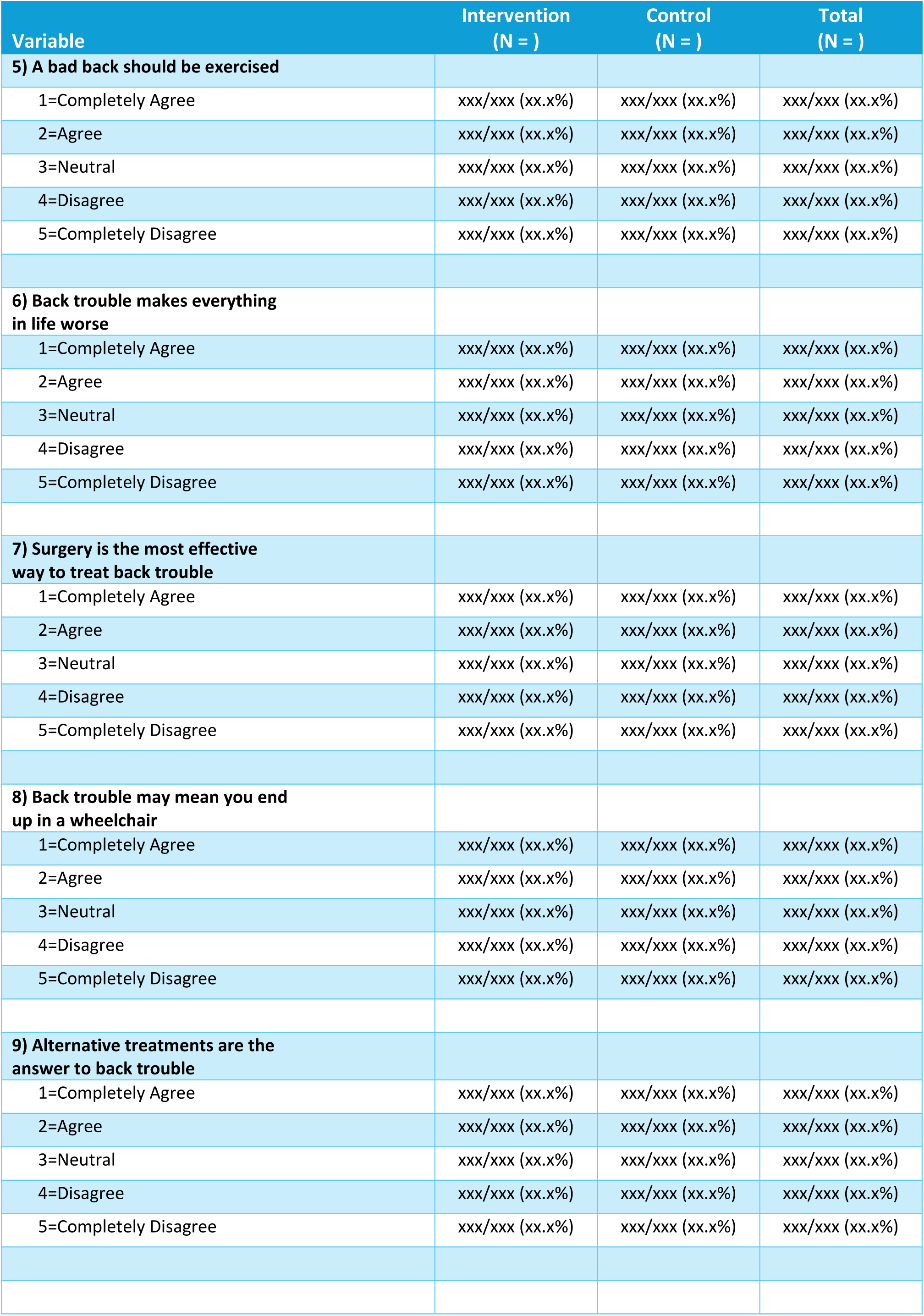
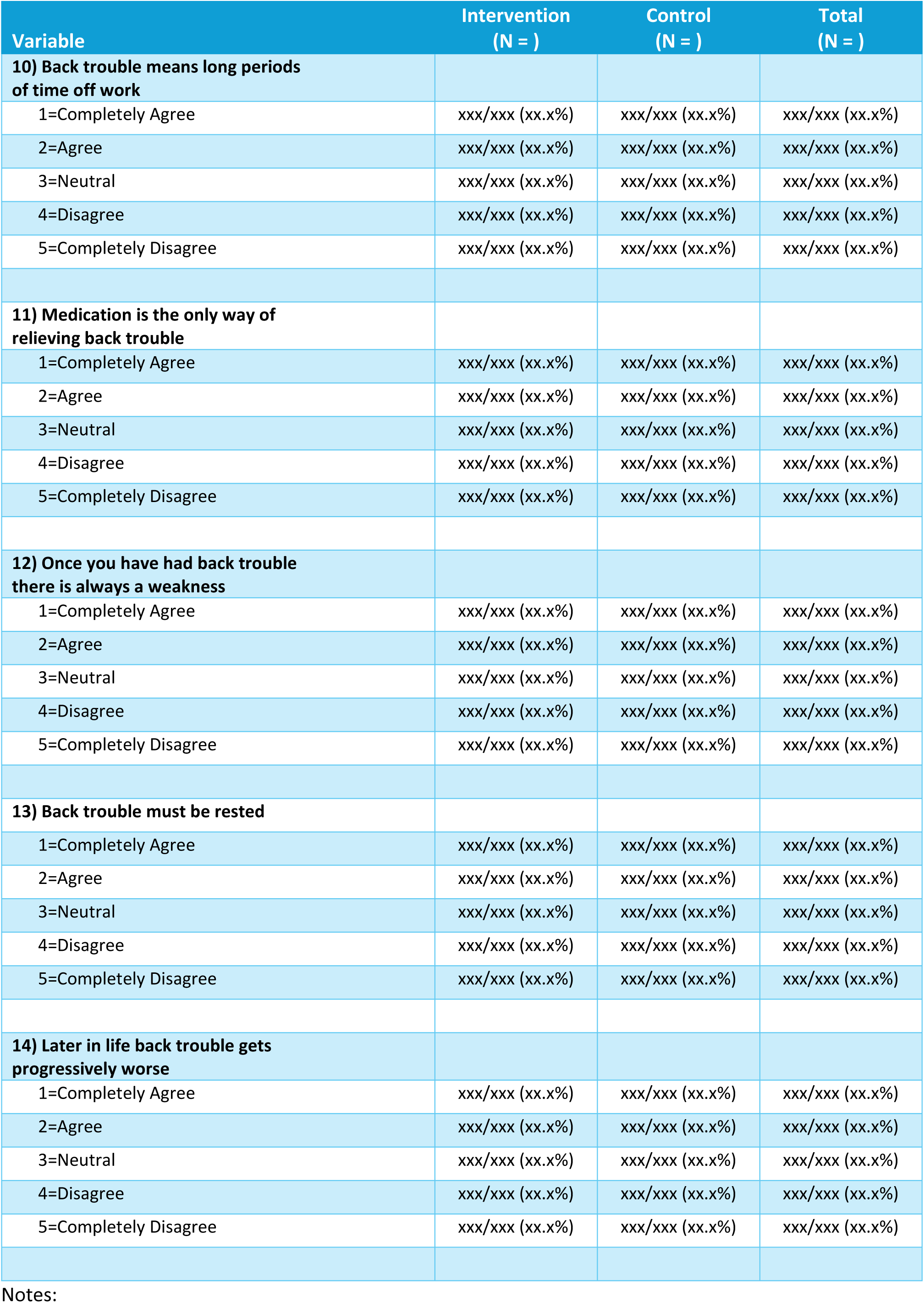
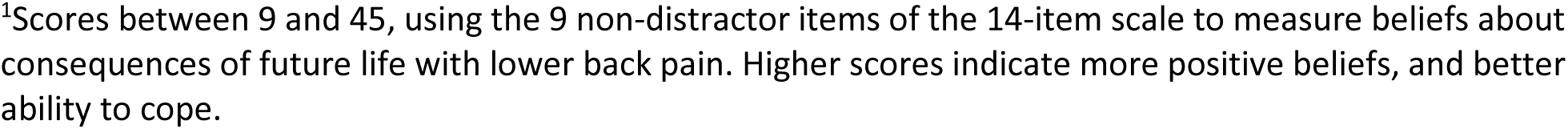
Participant-level outcome measure: The Back Beliefs Questionnaire (BBQ) results.

| Variable | Intervention<br>(N = ) | Control<br>(N = ) | Total<br>(N = ) |
| --- | --- | --- | --- |
| **Overall BBQ score<sup>1</sup>** |  |  |  |
| N | xxx | xxx | xxx |
| Mean (SD) | xx.x (xx.x) | xx.x (xx.x) | xx.x (xx.x) |
| Median (Q1; Q3) | xx.x (xx.x; xx.x) | xxx (xxx; xxx) | xxx (xxx; xxx) |
| Min Max | xxx xxx | xxx xxx | xxx xxx |
| **1) There is no real treatment for back trouble** |  |  |  |
| 1=Completely Agree | xxx/xxx (xx.x%) | xxx/xxx (xx.x%) | xxx/xxx (xx.x%) |
| 2=Agree | xxx/xxx (xx.x%) | xxx/xxx (xx.x%) | xxx/xxx (xx.x%) |
| 3=Neutral | xxx/xxx (xx.x%) | xxx/xxx (xx.x%) | xxx/xxx (xx.x%) |
| 4=Disagree | xxx/xxx (xx.x%) | xxx/xxx (xx.x%) | xxx/xxx (xx.x%) |
| 5=Completely Disagree | xxx/xxx (xx.x%) | xxx/xxx (xx.x%) | xxx/xxx (xx.x%) |
| **2) Back trouble will eventually stop you from working** |  |  |  |
| 1=Completely Agree | xxx/xxx (xx.x%) | xxx/xxx (xx.x%) | xxx/xxx (xx.x%) |
| 2=Agree | xxx/xxx (xx.x%) | xxx/xxx (xx.x%) | xxx/xxx (xx.x%) |
| 3=Neutral | xxx/xxx (xx.x%) | xxx/xxx (xx.x%) | xxx/xxx (xx.x%) |
| 4=Disagree | xxx/xxx (xx.x%) | xxx/xxx (xx.x%) | xxx/xxx (xx.x%) |
| 5=Completely Disagree | xxx/xxx (xx.x%) | xxx/xxx (xx.x%) | xxx/xxx (xx.x%) |
| **3) Back trouble means periods of pain for the rest of one's life** |  |  |  |
| 1=Completely Agree | xxx/xxx (xx.x%) | xxx/xxx (xx.x%) | xxx/xxx (xx.x%) |
| 2=Agree | xxx/xxx (xx.x%) | xxx/xxx (xx.x%) | xxx/xxx (xx.x%) |
| 3=Neutral | xxx/xxx (xx.x%) | xxx/xxx (xx.x%) | xxx/xxx (xx.x%) |
| 4=Disagree | xxx/xxx (xx.x%) | xxx/xxx (xx.x%) | xxx/xxx (xx.x%) |
| 5=Completely Disagree | xxx/xxx (xx.x%) | xxx/xxx (xx.x%) | xxx/xxx (xx.x%) |
| **4) Doctors cannot do anything for back trouble** |  |  |  |
| 1=Completely Agree | xxx/xxx (xx.x%) | xxx/xxx (xx.x%) | xxx/xxx (xx.x%) |
| 2=Agree | xxx/xxx (xx.x%) | xxx/xxx (xx.x%) | xxx/xxx (xx.x%) |
| 3=Neutral | xxx/xxx (xx.x%) | xxx/xxx (xx.x%) | xxx/xxx (xx.x%) |
| 4=Disagree | xxx/xxx (xx.x%) | xxx/xxx (xx.x%) | xxx/xxx (xx.x%) |
| 5=Completely Disagree | xxx/xxx (xx.x%) | xxx/xxx (xx.x%) | xxx/xxx (xx.x%) |
| **5) A bad back should be exercised** |  |  |  |
| 1=Completely Agree | xxx/xxx (xx.x%) | xxx/xxx (xx.x%) | xxx/xxx (xx.x%) |
| 2=Agree | xxx/xxx (xx.x%) | xxx/xxx (xx.x%) | xxx/xxx (xx.x%) |
| 3=Neutral | xxx/xxx (xx.x%) | xxx/xxx (xx.x%) | xxx/xxx (xx.x%) |
| 4=Disagree | xxx/xxx (xx.x%) | xxx/xxx (xx.x%) | xxx/xxx (xx.x%) |
| 5=Completely Disagree | xxx/xxx (xx.x%) | xxx/xxx (xx.x%) | xxx/xxx (xx.x%) |
| **6) Back trouble makes everything in life worse** |  |  |  |
| 1=Completely Agree | xxx/xxx (xx.x%) | xxx/xxx (xx.x%) | xxx/xxx (xx.x%) |
| 2=Agree | xxx/xxx (xx.x%) | xxx/xxx (xx.x%) | xxx/xxx (xx.x%) |
| 3=Neutral | xxx/xxx (xx.x%) | xxx/xxx (xx.x%) | xxx/xxx (xx.x%) |
| 4=Disagree | xxx/xxx (xx.x%) | xxx/xxx (xx.x%) | xxx/xxx (xx.x%) |
| 5=Completely Disagree | xxx/xxx (xx.x%) | xxx/xxx (xx.x%) | xxx/xxx (xx.x%) |
| **7) Surgery is the most effective way to treat back trouble** |  |  |  |
| 1=Completely Agree | xxx/xxx (xx.x%) | xxx/xxx (xx.x%) | xxx/xxx (xx.x%) |
| 2=Agree | xxx/xxx (xx.x%) | xxx/xxx (xx.x%) | xxx/xxx (xx.x%) |
| 3=Neutral | xxx/xxx (xx.x%) | xxx/xxx (xx.x%) | xxx/xxx (xx.x%) |
| 4=Disagree | xxx/xxx (xx.x%) | xxx/xxx (xx.x%) | xxx/xxx (xx.x%) |
| 5=Completely Disagree | xxx/xxx (xx.x%) | xxx/xxx (xx.x%) | xxx/xxx (xx.x%) |
| **8) Back trouble may mean you end up in a wheelchair** |  |  |  |
| 1=Completely Agree | xxx/xxx (xx.x%) | xxx/xxx (xx.x%) | xxx/xxx (xx.x%) |
| 2=Agree | xxx/xxx (xx.x%) | xxx/xxx (xx.x%) | xxx/xxx (xx.x%) |
| 3=Neutral | xxx/xxx (xx.x%) | xxx/xxx (xx.x%) | xxx/xxx (xx.x%) |
| 4=Disagree | xxx/xxx (xx.x%) | xxx/xxx (xx.x%) | xxx/xxx (xx.x%) |
| 5=Completely Disagree | xxx/xxx (xx.x%) | xxx/xxx (xx.x%) | xxx/xxx (xx.x%) |
| **9) Alternative treatments are the answer to back trouble** |  |  |  |
| 1=Completely Agree | xxx/xxx (xx.x%) | xxx/xxx (xx.x%) | xxx/xxx (xx.x%) |
| 2=Agree | xxx/xxx (xx.x%) | xxx/xxx (xx.x%) | xxx/xxx (xx.x%) |
| 3=Neutral | xxx/xxx (xx.x%) | xxx/xxx (xx.x%) | xxx/xxx (xx.x%) |
| 4=Disagree | xxx/xxx (xx.x%) | xxx/xxx (xx.x%) | xxx/xxx (xx.x%) |
| 5=Completely Disagree | xxx/xxx (xx.x%) | xxx/xxx (xx.x%) | xxx/xxx (xx.x%) |
| **10) Back trouble means long periods of time off work** |  |  |  |
| 1=Completely Agree | xxx/xxx (xx.x%) | xxx/xxx (xx.x%) | xxx/xxx (xx.x%) |
| 2=Agree | xxx/xxx (xx.x%) | xxx/xxx (xx.x%) | xxx/xxx (xx.x%) |
| 3=Neutral | xxx/xxx (xx.x%) | xxx/xxx (xx.x%) | xxx/xxx (xx.x%) |
| 4=Disagree | xxx/xxx (xx.x%) | xxx/xxx (xx.x%) | xxx/xxx (xx.x%) |
| 5=Completely Disagree | xxx/xxx (xx.x%) | xxx/xxx (xx.x%) | xxx/xxx (xx.x%) |
| **11) Medication is the only way of relieving back trouble** |  |  |  |
| 1=Completely Agree | xxx/xxx (xx.x%) | xxx/xxx (xx.x%) | xxx/xxx (xx.x%) |
| 2=Agree | xxx/xxx (xx.x%) | xxx/xxx (xx.x%) | xxx/xxx (xx.x%) |
| 3=Neutral | xxx/xxx (xx.x%) | xxx/xxx (xx.x%) | xxx/xxx (xx.x%) |
| 4=Disagree | xxx/xxx (xx.x%) | xxx/xxx (xx.x%) | xxx/xxx (xx.x%) |
| 5=Completely Disagree | xxx/xxx (xx.x%) | xxx/xxx (xx.x%) | xxx/xxx (xx.x%) |
| **12) Once you have had back trouble there is always a weakness** |  |  |  |
| 1=Completely Agree | xxx/xxx (xx.x%) | xxx/xxx (xx.x%) | xxx/xxx (xx.x%) |
| 2=Agree | xxx/xxx (xx.x%) | xxx/xxx (xx.x%) | xxx/xxx (xx.x%) |
| 3=Neutral | xxx/xxx (xx.x%) | xxx/xxx (xx.x%) | xxx/xxx (xx.x%) |
| 4=Disagree | xxx/xxx (xx.x%) | xxx/xxx (xx.x%) | xxx/xxx (xx.x%) |
| 5=Completely Disagree | xxx/xxx (xx.x%) | xxx/xxx (xx.x%) | xxx/xxx (xx.x%) |
| **13) Back trouble must be rested** |  |  |  |
| 1=Completely Agree | xxx/xxx (xx.x%) | xxx/xxx (xx.x%) | xxx/xxx (xx.x%) |
| 2=Agree | xxx/xxx (xx.x%) | xxx/xxx (xx.x%) | xxx/xxx (xx.x%) |
| 3=Neutral | xxx/xxx (xx.x%) | xxx/xxx (xx.x%) | xxx/xxx (xx.x%) |
| 4=Disagree | xxx/xxx (xx.x%) | xxx/xxx (xx.x%) | xxx/xxx (xx.x%) |
| 5=Completely Disagree | xxx/xxx (xx.x%) | xxx/xxx (xx.x%) | xxx/xxx (xx.x%) |
| **14) Later in life back trouble gets progressively worse** |  |  |  |
| 1=Completely Agree | xxx/xxx (xx.x%) | xxx/xxx (xx.x%) | xxx/xxx (xx.x%) |
| 2=Agree | xxx/xxx (xx.x%) | xxx/xxx (xx.x%) | xxx/xxx (xx.x%) |
| 3=Neutral | xxx/xxx (xx.x%) | xxx/xxx (xx.x%) | xxx/xxx (xx.x%) |
| 4=Disagree | xxx/xxx (xx.x%) | xxx/xxx (xx.x%) | xxx/xxx (xx.x%) |
| 5=Completely Disagree | xxx/xxx (xx.x%) | xxx/xxx (xx.x%) | xxx/xxx (xx.x%) |

Notes:
<sup>1</sup>Scores between 9 and 45, using the 9 non-distractor items of the 14-item scale to measure beliefs about consequences of future life with lower back pain. Higher scores indicate more positive beliefs, and better ability to cope.

**Table 5.** Participant-level outcome measure: The Back Pain Attitude Questionnaire (Back-PAQ) results.

| Variable | Intervention<br>(N = ) | Control<br>(N = ) | Total<br>(N = ) |
| --- | --- | --- | --- |
| **Overall Back-PAQ score<sup>1</sup>** |  |  |  |
| N | xxx | xxx | xxx |
| Mean (SD) | xx.x (xx.x) | xx.x (xx.x) | xx.x (xx.x) |
| Median (Q1; Q3) | xx.x (xx.x; xx.x) | xxx (xxx; xxx) | xxx (xxx; xxx) |
| Min Max | xxx xxx | xxx xxx | xxx xxx |
| **THE PATIENT'S OWN BACK** |  |  |  |
| **1) Bending your back is good for it** |  |  |  |
| 1=False | xxx/xxx (xx.x%) | xxx/xxx (xx.x%) | xxx/xxx (xx.x%) |
| 2=Possibly False | xxx/xxx (xx.x%) | xxx/xxx (xx.x%) | xxx/xxx (xx.x%) |
| 3=Unsure | xxx/xxx (xx.x%) | xxx/xxx (xx.x%) | xxx/xxx (xx.x%) |
| 4=Possibly True | xxx/xxx (xx.x%) | xxx/xxx (xx.x%) | xxx/xxx (xx.x%) |
| 5=True | xxx/xxx (xx.x%) | xxx/xxx (xx.x%) | xxx/xxx (xx.x%) |
| **2) It is easy to injure your back** |  |  |  |
| 1=False | xxx/xxx (xx.x%) | xxx/xxx (xx.x%) | xxx/xxx (xx.x%) |
| 2=Possibly False | xxx/xxx (xx.x%) | xxx/xxx (xx.x%) | xxx/xxx (xx.x%) |
| 3=Unsure | xxx/xxx (xx.x%) | xxx/xxx (xx.x%) | xxx/xxx (xx.x%) |
| 4=Possibly True | xxx/xxx (xx.x%) | xxx/xxx (xx.x%) | xxx/xxx (xx.x%) |
| 5=True | xxx/xxx (xx.x%) | xxx/xxx (xx.x%) | xxx/xxx (xx.x%) |
| **3) If you overuse your back, it will wear out** |  |  |  |
| 1=False | xxx/xxx (xx.x%) | xxx/xxx (xx.x%) | xxx/xxx (xx.x%) |
| 2=Possibly False | xxx/xxx (xx.x%) | xxx/xxx (xx.x%) | xxx/xxx (xx.x%) |
| 3=Unsure | xxx/xxx (xx.x%) | xxx/xxx (xx.x%) | xxx/xxx (xx.x%) |
| 4=Possibly True | xxx/xxx (xx.x%) | xxx/xxx (xx.x%) | xxx/xxx (xx.x%) |
| 5=True | xxx/xxx (xx.x%) | xxx/xxx (xx.x%) | xxx/xxx (xx.x%) |
| **4) If an activity or movement causes back pain, you should avoid it in the future** |  |  |  |
| 1=False | xxx/xxx (xx.x%) | xxx/xxx (xx.x%) | xxx/xxx (xx.x%) |
| 2=Possibly False | xxx/xxx (xx.x%) | xxx/xxx (xx.x%) | xxx/xxx (xx.x%) |
| 3=Unsure | xxx/xxx (xx.x%) | xxx/xxx (xx.x%) | xxx/xxx (xx.x%) |
| 4=Possibly True | xxx/xxx (xx.x%) | xxx/xxx (xx.x%) | xxx/xxx (xx.x%) |
| 5=True | xxx/xxx (xx.x%) | xxx/xxx (xx.x%) | xxx/xxx (xx.x%) |
| **5) You could injure your back if you are not careful** |  |  |  |
| 1=False | xxx/xxx (xx.x%) | xxx/xxx (xx.x%) | xxx/xxx (xx.x%) |
| 2=Possibly False | xxx/xxx (xx.x%) | xxx/xxx (xx.x%) | xxx/xxx (xx.x%) |
| 3=Unsure | xxx/xxx (xx.x%) | xxx/xxx (xx.x%) | xxx/xxx (xx.x%) |
| 4=Possibly True | xxx/xxx (xx.x%) | xxx/xxx (xx.x%) | xxx/xxx (xx.x%) |
| 5=True | xxx/xxx (xx.x%) | xxx/xxx (xx.x%) | xxx/xxx (xx.x%) |
| **6) Back pain means that you have injured your back** |  |  |  |
| 1=False | xxx/xxx (xx.x%) | xxx/xxx (xx.x%) | xxx/xxx (xx.x%) |
| 2=Possibly False | xxx/xxx (xx.x%) | xxx/xxx (xx.x%) | xxx/xxx (xx.x%) |
| 3=Unsure | xxx/xxx (xx.x%) | xxx/xxx (xx.x%) | xxx/xxx (xx.x%) |
| 4=Possibly True | xxx/xxx (xx.x%) | xxx/xxx (xx.x%) | xxx/xxx (xx.x%) |
| 5=True | xxx/xxx (xx.x%) | xxx/xxx (xx.x%) | xxx/xxx (xx.x%) |
| **7) A twinge in your back can be the first sign of a serious injury** |  |  |  |
| 1=False | xxx/xxx (xx.x%) | xxx/xxx (xx.x%) | xxx/xxx (xx.x%) |
| 2=Possibly False | xxx/xxx (xx.x%) | xxx/xxx (xx.x%) | xxx/xxx (xx.x%) |
| 3=Unsure | xxx/xxx (xx.x%) | xxx/xxx (xx.x%) | xxx/xxx (xx.x%) |
| 4=Possibly True | xxx/xxx (xx.x%) | xxx/xxx (xx.x%) | xxx/xxx (xx.x%) |
| 5=True | xxx/xxx (xx.x%) | xxx/xxx (xx.x%) | xxx/xxx (xx.x%) |
| **8) Having back pain makes it difficult to enjoy life** |  |  |  |
| 1=False | xxx/xxx (xx.x%) | xxx/xxx (xx.x%) | xxx/xxx (xx.x%) |
| 2=Possibly False | xxx/xxx (xx.x%) | xxx/xxx (xx.x%) | xxx/xxx (xx.x%) |
| 3=Unsure | xxx/xxx (xx.x%) | xxx/xxx (xx.x%) | xxx/xxx (xx.x%) |
| 4=Possibly True | xxx/xxx (xx.x%) | xxx/xxx (xx.x%) | xxx/xxx (xx.x%) |
| 5=True | xxx/xxx (xx.x%) | xxx/xxx (xx.x%) | xxx/xxx (xx.x%) |
| **9) It is worse to have pain in your back than your arms or legs** |  |  |  |
| 1=False | xxx/xxx (xx.x%) | xxx/xxx (xx.x%) | xxx/xxx (xx.x%) |
| 2=Possibly False | xxx/xxx (xx.x%) | xxx/xxx (xx.x%) | xxx/xxx (xx.x%) |
| 3=Unsure | xxx/xxx (xx.x%) | xxx/xxx (xx.x%) | xxx/xxx (xx.x%) |
| 4=Possibly True | xxx/xxx (xx.x%) | xxx/xxx (xx.x%) | xxx/xxx (xx.x%) |
| 5=True | xxx/xxx (xx.x%) | xxx/xxx (xx.x%) | xxx/xxx (xx.x%) |
| **10) It is hard to understand what back pain is like if you have never had it yourself** |  |  |  |
| 1=False | xxx/xxx (xx.x%) | xxx/xxx (xx.x%) | xxx/xxx (xx.x%) |
| 2=Possibly False | xxx/xxx (xx.x%) | xxx/xxx (xx.x%) | xxx/xxx (xx.x%) |
| 3=Unsure | xxx/xxx (xx.x%) | xxx/xxx (xx.x%) | xxx/xxx (xx.x%) |
| 4=Possibly True | xxx/xxx (xx.x%) | xxx/xxx (xx.x%) | xxx/xxx (xx.x%) |
| 5=True | xxx/xxx (xx.x%) | xxx/xxx (xx.x%) | xxx/xxx (xx.x%) |
| **RECOVERING FROM BACK PAIN** |  |  |  |
| **11) If your back hurts, you should take it easy until the pain goes away** |  |  |  |
| 1=False | xxx/xxx (xx.x%) | xxx/xxx (xx.x%) | xxx/xxx (xx.x%) |
| 2=Possibly False | xxx/xxx (xx.x%) | xxx/xxx (xx.x%) | xxx/xxx (xx.x%) |
| 3=Unsure | xxx/xxx (xx.x%) | xxx/xxx (xx.x%) | xxx/xxx (xx.x%) |
| 4=Possibly True | xxx/xxx (xx.x%) | xxx/xxx (xx.x%) | xxx/xxx (xx.x%) |
| 5=True | xxx/xxx (xx.x%) | xxx/xxx (xx.x%) | xxx/xxx (xx.x%) |
| **12) If you ignore back pain, you may cause damage to your back** |  |  |  |
| 1=False | xxx/xxx (xx.x%) | xxx/xxx (xx.x%) | xxx/xxx (xx.x%) |
| 2=Possibly False | xxx/xxx (xx.x%) | xxx/xxx (xx.x%) | xxx/xxx (xx.x%) |
| 3=Unsure | xxx/xxx (xx.x%) | xxx/xxx (xx.x%) | xxx/xxx (xx.x%) |
| 4=Possibly True | xxx/xxx (xx.x%) | xxx/xxx (xx.x%) | xxx/xxx (xx.x%) |
| 5=True | xxx/xxx (xx.x%) | xxx/xxx (xx.x%) | xxx/xxx (xx.x%) |
| **13) It is important to see a health professional when you have back pain** |  |  |  |
| 1=False | xxx/xxx (xx.x%) | xxx/xxx (xx.x%) | xxx/xxx (xx.x%) |
| 2=Possibly False | xxx/xxx (xx.x%) | xxx/xxx (xx.x%) | xxx/xxx (xx.x%) |
| 3=Unsure | xxx/xxx (xx.x%) | xxx/xxx (xx.x%) | xxx/xxx (xx.x%) |
| 4=Possibly True | xxx/xxx (xx.x%) | xxx/xxx (xx.x%) | xxx/xxx (xx.x%) |
| 5=True | xxx/xxx (xx.x%) | xxx/xxx (xx.x%) | xxx/xxx (xx.x%) |
| **14) To effectively treat back pain you need to know exactly what is wrong** |  |  |  |
| 1=False | xxx/xxx (xx.x%) | xxx/xxx (xx.x%) | xxx/xxx (xx.x%) |
| 2=Possibly False | xxx/xxx (xx.x%) | xxx/xxx (xx.x%) | xxx/xxx (xx.x%) |
| 3=Unsure | xxx/xxx (xx.x%) | xxx/xxx (xx.x%) | xxx/xxx (xx.x%) |
| 4=Possibly True | xxx/xxx (xx.x%) | xxx/xxx (xx.x%) | xxx/xxx (xx.x%) |
| 5=True | xxx/xxx (xx.x%) | xxx/xxx (xx.x%) | xxx/xxx (xx.x%) |
| **15) If you have back pain you should avoid exercise** |  |  |  |
| 1=False | xxx/xxx (xx.x%) | xxx/xxx (xx.x%) | xxx/xxx (xx.x%) |
| 2=Possibly False | xxx/xxx (xx.x%) | xxx/xxx (xx.x%) | xxx/xxx (xx.x%) |
| 3=Unsure | xxx/xxx (xx.x%) | xxx/xxx (xx.x%) | xxx/xxx (xx.x%) |
| 4=Possibly True | xxx/xxx (xx.x%) | xxx/xxx (xx.x%) | xxx/xxx (xx.x%) |
| 5=True | xxx/xxx (xx.x%) | xxx/xxx (xx.x%) | xxx/xxx (xx.x%) |
| **16) When you have back pain the risks of vigorous exercise outweigh the benefits** |  |  |  |
| 1=False | xxx/xxx (xx.x%) | xxx/xxx (xx.x%) | xxx/xxx (xx.x%) |
| 2=Possibly False | xxx/xxx (xx.x%) | xxx/xxx (xx.x%) | xxx/xxx (xx.x%) |
| 3=Unsure | xxx/xxx (xx.x%) | xxx/xxx (xx.x%) | xxx/xxx (xx.x%) |
| 4=Possibly True | xxx/xxx (xx.x%) | xxx/xxx (xx.x%) | xxx/xxx (xx.x%) |
| 5=True | xxx/xxx (xx.x%) | xxx/xxx (xx.x%) | xxx/xxx (xx.x%) |
| **17) If you have back pain you should try to stay active** |  |  |  |
| 1=False | xxx/xxx (xx.x%) | xxx/xxx (xx.x%) | xxx/xxx (xx.x%) |
| 2=Possibly False | xxx/xxx (xx.x%) | xxx/xxx (xx.x%) | xxx/xxx (xx.x%) |
| 3=Unsure | xxx/xxx (xx.x%) | xxx/xxx (xx.x%) | xxx/xxx (xx.x%) |
| 4=Possibly True | xxx/xxx (xx.x%) | xxx/xxx (xx.x%) | xxx/xxx (xx.x%) |
| 5=True | xxx/xxx (xx.x%) | xxx/xxx (xx.x%) | xxx/xxx (xx.x%) |
| **18) Once you have had back pain there is always a weakness** |  |  |  |
| 1=False | xxx/xxx (xx.x%) | xxx/xxx (xx.x%) | xxx/xxx (xx.x%) |
| 2=Possibly False | xxx/xxx (xx.x%) | xxx/xxx (xx.x%) | xxx/xxx (xx.x%) |
| 3=Unsure | xxx/xxx (xx.x%) | xxx/xxx (xx.x%) | xxx/xxx (xx.x%) |
| 4=Possibly True | xxx/xxx (xx.x%) | xxx/xxx (xx.x%) | xxx/xxx (xx.x%) |
| 5=True | xxx/xxx (xx.x%) | xxx/xxx (xx.x%) | xxx/xxx (xx.x%) |
| **19) There is a high chance that an episode of back pain will not resolve** |  |  |  |
| 1=False | xxx/xxx (xx.x%) | xxx/xxx (xx.x%) | xxx/xxx (xx.x%) |
| 2=Possibly False | xxx/xxx (xx.x%) | xxx/xxx (xx.x%) | xxx/xxx (xx.x%) |
| 3=Unsure | xxx/xxx (xx.x%) | xxx/xxx (xx.x%) | xxx/xxx (xx.x%) |
| 4=Possibly True | xxx/xxx (xx.x%) | xxx/xxx (xx.x%) | xxx/xxx (xx.x%) |
| 5=True | xxx/xxx (xx.x%) | xxx/xxx (xx.x%) | xxx/xxx (xx.x%) |
| **20) Once you have a back problem, there is not a lot you can do about it** |  |  |  |
| 1=False | xxx/xxx (xx.x%) | xxx/xxx (xx.x%) | xxx/xxx (xx.x%) |
| 2=Possibly False | xxx/xxx (xx.x%) | xxx/xxx (xx.x%) | xxx/xxx (xx.x%) |
| 3=Unsure | xxx/xxx (xx.x%) | xxx/xxx (xx.x%) | xxx/xxx (xx.x%) |
| 4=Possibly True | xxx/xxx (xx.x%) | xxx/xxx (xx.x%) | xxx/xxx (xx.x%) |
| 5=True | xxx/xxx (xx.x%) | xxx/xxx (xx.x%) | xxx/xxx (xx.x%) |

Notes:
<sup>1</sup>Scores between 20 and 100, of 20-item scale to evaluate attitudes and beliefs of the public, people with back pain, and healthcare professionals about the spine. Higher scores indicate more unhelpful beliefs.

**Table 6.** Participant Video Feedback Survey results.

| Variable | Intervention<br>(N = ) | Control<br>(N = ) | Total<br>(N = ) |
| --- | --- | --- | --- |
| **The participant's view of the video intervention being helpful to support the self-management of their low back pain** |  |  |  |
| Strongly disagree | xxx/xxx (xx.x%) | xxx/xxx (xx.x%) | xxx/xxx (xx.x%) |
| Disagree | xxx/xxx (xx.x%) | xxx/xxx (xx.x%) | xxx/xxx (xx.x%) |
| Neutral | xxx/xxx (xx.x%) | xxx/xxx (xx.x%) | xxx/xxx (xx.x%) |
| Agree | xxx/xxx (xx.x%) | xxx/xxx (xx.x%) | xxx/xxx (xx.x%) |
| Strongly agree | xxx/xxx (xx.x%) | xxx/xxx (xx.x%) | xxx/xxx (xx.x%) |
| **The participant's view of the effectiveness of the video intervention** |  |  |  |
| Yes very effective | xxx/xxx (xx.x%) | xxx/xxx (xx.x%) | xxx/xxx (xx.x%) |
| Somewhat effective | xxx/xxx (xx.x%) | xxx/xxx (xx.x%) | xxx/xxx (xx.x%) |
| Neutral | xxx/xxx (xx.x%) | xxx/xxx (xx.x%) | xxx/xxx (xx.x%) |
| Somewhat ineffective | xxx/xxx (xx.x%) | xxx/xxx (xx.x%) | xxx/xxx (xx.x%) |
| Very ineffective | xxx/xxx (xx.x%) | xxx/xxx (xx.x%) | xxx/xxx (xx.x%) |
| **The participant's view of their engagement with the video intervention** |  |  |  |
| Strongly disagree | xxx/xxx (xx.x%) | xxx/xxx (xx.x%) | xxx/xxx (xx.x%) |
| Disagree | xxx/xxx (xx.x%) | xxx/xxx (xx.x%) | xxx/xxx (xx.x%) |
| Neutral | xxx/xxx (xx.x%) | xxx/xxx (xx.x%) | xxx/xxx (xx.x%) |
| Agree | xxx/xxx (xx.x%) | xxx/xxx (xx.x%) | xxx/xxx (xx.x%) |
| Strongly agree | xxx/xxx (xx.x%) | xxx/xxx (xx.x%) | xxx/xxx (xx.x%) |
| **The participant's view of whether the video intervention can be improved<sup>1</sup>** |  |  |  |
| No improvement is required | xxx/xxx (xx.x%) | xxx/xxx (xx.x%) | xxx/xxx (xx.x%) |
| Yes, the video could be longer | xxx/xxx (xx.x%) | xxx/xxx (xx.x%) | xxx/xxx (xx.x%) |
| Yes, the video could be played more often | xxx/xxx (xx.x%) | xxx/xxx (xx.x%) | xxx/xxx (xx.x%) |
| Yes, the video's information could be clearer | xxx/xxx (xx.x%) | xxx/xxx (xx.x%) | xxx/xxx (xx.x%) |
| Yes, the video's language could be better | xxx/xxx (xx.x%) | xxx/xxx (xx.x%) | xxx/xxx (xx.x%) |
| Yes, others | xxx/xxx (xx.x%) | xxx/xxx (xx.x%) | xxx/xxx (xx.x%) |
| **How the participant views the video intervention can be improved further** |  |  |  |
| {list responses} | xxx/xxx (xx.x%) | xxx/xxx (xx.x%) | xxx/xxx (xx.x%) |
| **The participant's view of whether the video intervention changed their beliefs about their low back pain** |  |  |  |
| Strongly disagree | xxx/xxx (xx.x%) | xxx/xxx (xx.x%) | xxx/xxx (xx.x%) |
| Disagree | xxx/xxx (xx.x%) | xxx/xxx (xx.x%) | xxx/xxx (xx.x%) |
| Neutral | xxx/xxx (xx.x%) | xxx/xxx (xx.x%) | xxx/xxx (xx.x%) |
| Agree | xxx/xxx (xx.x%) | xxx/xxx (xx.x%) | xxx/xxx (xx.x%) |
| Strongly agree | xxx/xxx (xx.x%) | xxx/xxx (xx.x%) | xxx/xxx (xx.x%) |
| **How the participant views the video intervention changed their beliefs about their low back pain** |  |  |  |
| {list responses} | xxx/xxx (xx.x%) | xxx/xxx (xx.x%) | xxx/xxx (xx.x%) |
| **The participant's view of the influence of the video intervention on the way they interact with their health providers** |  |  |  |
| Strongly disagree | xxx/xxx (xx.x%) | xxx/xxx (xx.x%) | xxx/xxx (xx.x%) |
| Disagree | xxx/xxx (xx.x%) | xxx/xxx (xx.x%) | xxx/xxx (xx.x%) |
| Neutral | xxx/xxx (xx.x%) | xxx/xxx (xx.x%) | xxx/xxx (xx.x%) |
| Agree | xxx/xxx (xx.x%) | xxx/xxx (xx.x%) | xxx/xxx (xx.x%) |
| Strongly agree | xxx/xxx (xx.x%) | xxx/xxx (xx.x%) | xxx/xxx (xx.x%) |
| **How the participant views the video intervention has influenced the way they interact with their health providers** |  |  |  |
| {list responses} | xxx/xxx (xx.x%) | xxx/xxx (xx.x%) | xxx/xxx (xx.x%) |
| **How the participant believes the video can be used to assist people's recovery from low back pain** |  |  |  |
| {list responses} | xxx/xxx (xx.x%) | xxx/xxx (xx.x%) | xxx/xxx (xx.x%) |

Notes:
<sup>1</sup>Multiple response categories may be selected by the participant

**Table 7.**
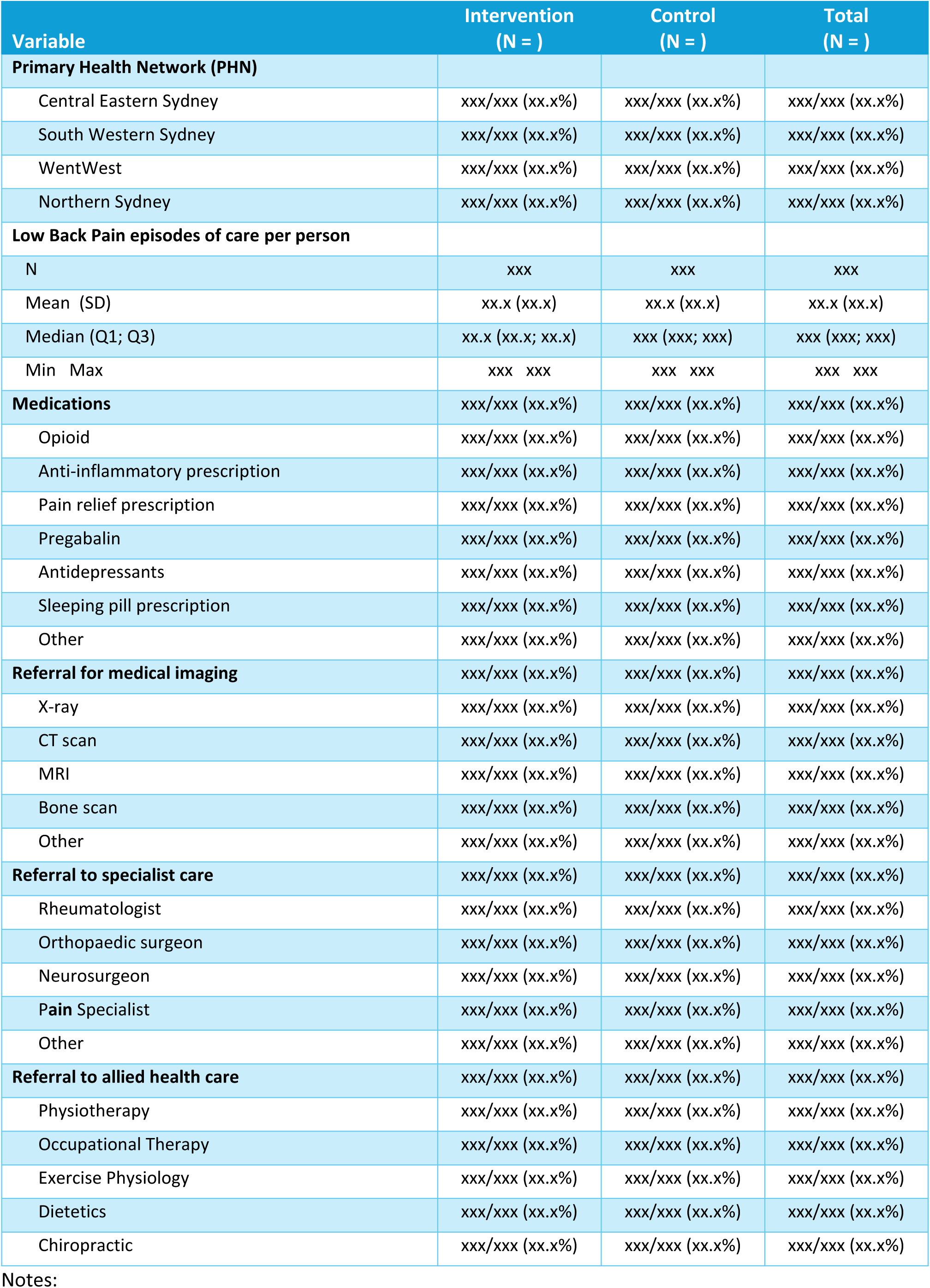
Practice-level descriptive results.

**Table 8.** Participant-level model outcomes.

| Outcome | Unadjusted model <sup>1</sup> |  |  |  |  | Adjusted model <sup>2</sup> |  |  |  |  |
| --- | --- | --- | --- | --- | --- | --- | --- | --- | --- | --- |
|  | N | MD (95% CI) | P | P-adj | ICC | N | MD (95% CI) | P | P-adj | ICC |
| **The Back Beliefs Questionnaire (BBQ)** | xxxx | xx.xx (xx.xx to xx.xx) | 0.xxx | 0.xxx | 0.xx | xxxx | xx.xx (xx.xx to xx.xx) | 0.xxx | 0.xxx | 0.xx |
| **The Back Pain Attitude Questionnaire (Back-PAQ)** | xxxx | xx.xx (xx.xx to xx.xx) | 0.xxx | 0.xxx | 0.xx | xxxx | xx.xx (xx.xx to xx.xx) | 0.xxx | 0.xxx | 0.xx |

Notes:
- 1) For continuous and ordinal outcomes, the unadjusted model consists in a generalised linear mixed model with a random effect for cluster (GP clinic), a fixed effect indicating the group assignment of each cluster at each step, and a fixed categorical effect of time (each step). Appropriate distributions and link functions are used depending on the nature of the outcome. - 2) Adjusted models include the additional baseline covariates: age (continuous), sex (binary), presently have LBP (binary)
**MD** = Mean Difference; **CI** = Confidence Interval; **P** = p-value; **P-adj** = p-value after Holm- Bonferroni adjustment; **ICC** = intra-cluster correlation
All p-values are ordered from smallest to largest and tested in sequence. At each step, the p-value is compared with $\alpha = 1 - (1 - 0.05)^{**1/C}$ where C is the number of hypotheses remaining. With two outcomes, the critical threshold levels are 0.025, and 0.05 for the first and last tests, respectively.

**Table 9.** Practice-level model outcomes.

| Outcome | Unadjusted model <sup>1</sup> |  |  |  |  | Adjusted model <sup>2</sup> |  |  |  |  |
| --- | --- | --- | --- | --- | --- | --- | --- | --- | --- | --- |
|  | N | OR (95% CI) | P | P-adj | ICC | N | OR (95% CI) | P | P-adj | ICC |
| **Opioid Medication** | xxxx | xx.xx (xx.xx to xx.xx) | 0.xxx | 0.xxx | 0.xx | xxxx | xx.xx (xx.xx to xx.xx) | 0.xxx | 0.xxx | 0.xx |
| **Referral for any medical imaging** | xxxx | xx.xx (xx.xx to xx.xx) | 0.xxx | 0.xxx | 0.xx | xxxx | xx.xx (xx.xx to xx.xx) | 0.xxx | 0.xxx | 0.xx |
| **Referral to any allied health care** | xxxx | xx.xx (xx.xx to xx.xx) | 0.xxx | 0.xxx | 0.xx | xxxx | xx.xx (xx.xx to xx.xx) | 0.xxx | 0.xxx | 0.xx |

Notes:
- 1) For continuous and ordinal outcomes, the unadjusted model consists in a generalised linear mixed model with a random effect for cluster (GP clinic), a fixed effect indicating the group assignment of each cluster at each step, and a fixed categorical effect of time (each step). Appropriate distributions and link functions are used depending on the nature of the outcome. - 2) Adjusted models include the additional baseline covariates: age (continuous), sex (binary)
**OR** = Odds Ratio; **CI** = Confidence Interval; **P** = p-value; **P-adj** = p-value after Holm- Bonferroni adjustment; **ICC** = intra-cluster correlation
All p-values are ordered from smallest to largest and tested in sequence. At each step, the p-value is compared with $\alpha = 1 - (1 - 0.05)^{**1/C}$ where C is the number of hypotheses remaining. With three outcomes, the critical threshold levels are 0.0167, 0.025, and 0.05 for the first, second and last tests, respectively.

**Table 10.** Other practice-level outcomes.

| Outcome | N | OR (95% CI) |
| --- | --- | --- |
| **Medications** |  |  |
| Anti-inflammatory prescription | xxxx | xx.xx (xx.xx to xx.xx) |
| Pain relief prescription | xxxx | xx.xx (xx.xx to xx.xx) |
| Pregabalin | xxxx | xx.xx (xx.xx to xx.xx) |
| Antidepressants | xxxx | xx.xx (xx.xx to xx.xx) |
| Sleeping pill prescription | xxxx | xx.xx (xx.xx to xx.xx) |
| Other | xxxx | xx.xx (xx.xx to xx.xx) |
| **Referral for medical imaging** |  |  |
| X-ray | xxxx | xx.xx (xx.xx to xx.xx) |
| CT scan | xxxx | xx.xx (xx.xx to xx.xx) |
| MRI | xxxx | xx.xx (xx.xx to xx.xx) |
| Bone scan | xxxx | xx.xx (xx.xx to xx.xx) |
| Other | xxxx | xx.xx (xx.xx to xx.xx) |
| **Referral to specialist care** |  |  |
| Rheumatologist | xxxx | xx.xx (xx.xx to xx.xx) |
| Orthopaedic surgeon | xxxx | xx.xx (xx.xx to xx.xx) |
| Neurosurgeon | xxxx | xx.xx (xx.xx to xx.xx) |
| Pain Specialist | xxxx | xx.xx (xx.xx to xx.xx) |
| Other | xxxx | xx.xx (xx.xx to xx.xx) |
| **Referral to allied health care** |  |  |
| Physiotherapy | xxxx | xx.xx (xx.xx to xx.xx) |
| Occupational Therapy | xxxx | xx.xx (xx.xx to xx.xx) |
| Exercise Physiology | xxxx | xx.xx (xx.xx to xx.xx) |
| Dietetics | xxxx | xx.xx (xx.xx to xx.xx) |
| Chiropractic | xxxx | xx.xx (xx.xx to xx.xx) |

**Notes:**
- 1) For continuous and ordinal outcomes, the unadjusted model consists in a generalised linear mixed model with a random effect for cluster (GP clinic), a fixed effect indicating the group assignment of each cluster at each step, and a fixed categorical effect of time (each step). Appropriate distributions and link functions are used depending on the nature of the outcome.
**OR** = Odds Ratio; **CI** = Confidence Interval

### 5.2 Figures

**Figure 1.**
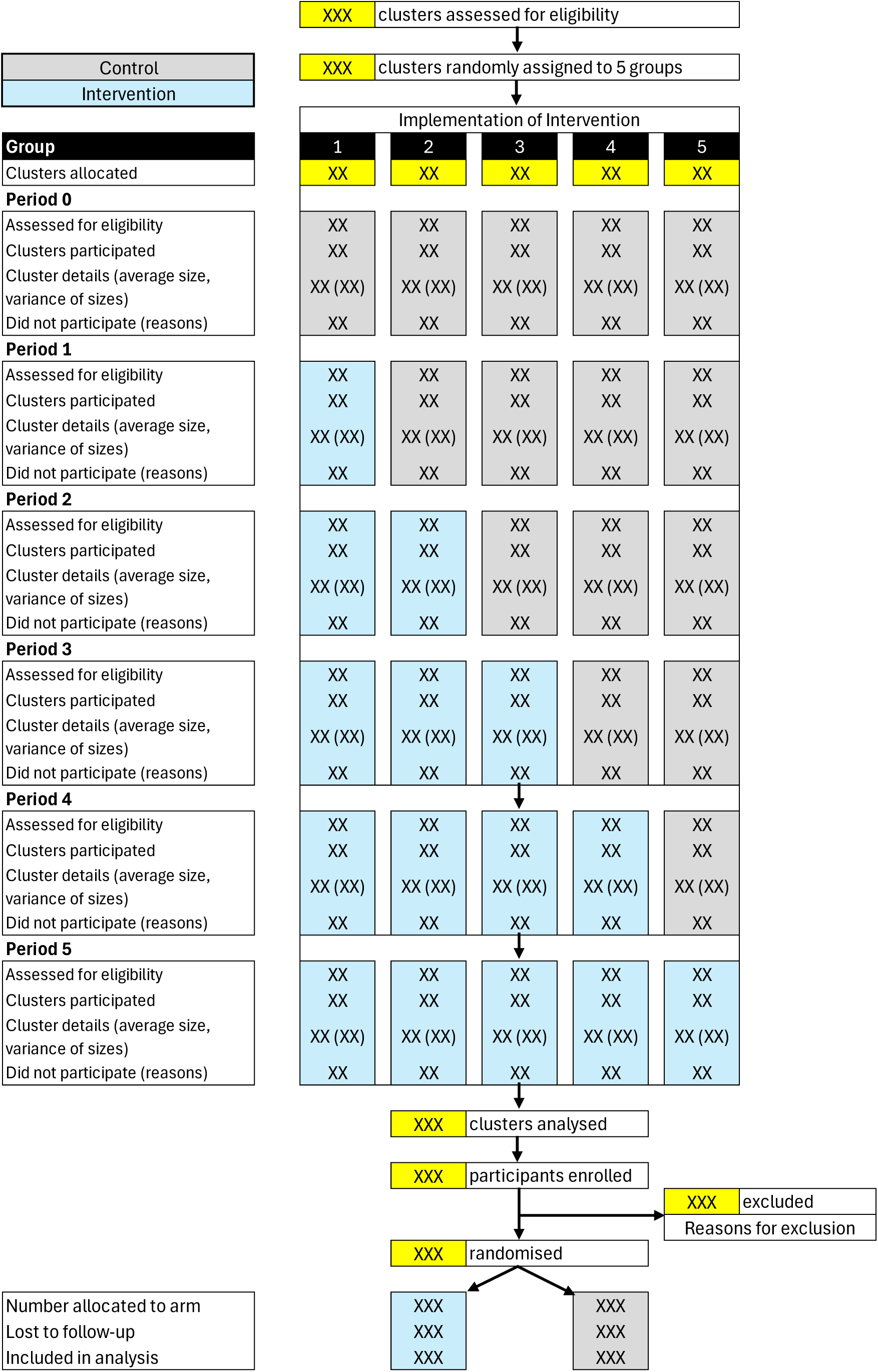
CONSORT diagram.

**Figure 2. Grotta bar charts of BBQ score**

Programming note: Stacked bar chart with 2 bars (intervention vs control) per question, grouped by non-distractor and distractor questions. Each bar to be of the same high (100%). Show the proportion in each category using labels on the bars.

**Figure 3. Grotta bar charts of Back-PAQ score**

Programming note: Stacked bar chart with 2 bars (intervention vs control) per question, grouped by questions about the patient’s own back, and questions about recovering from back pain’. Each bar to be of the same high (100%). Show the proportion in each category using labels on the bars.

**Figure 4. LBP episodes over time**

**Figure 5. Bar charts of LBP medications and referrals for imaging and care**

## Footnotes

1 As per the screening survey the participant will not be eligible to complete the survey again if they have previously completed one.

## References

1. Symonds, T.L., et al., Do attitudes and beliefs influence work loss due to low back trouble? Occup Med, 1996. 46(1): p. 25–32.

2. Krageloha, C., et al., Rasch analysis of the Back Pain Attitudes Questionnaire (Back-PAQ). Disabil Rehabil, 2022. 4(13): p. 3228–3235.

3. Hemming K, Taljaard M, McKenzie JE, Hooper R, Copas A, Thompson JA, et al. Reporting of stepped wedge cluster randomised trials: extension of the CONSORT 2010 statement with explanation and elaboration. BMJ 2018 Nov 9;363:k1614.

4. Holm, S. (1979) A Simple Sequentially Rejective Multiple Test Procedure. Scandinavian Journal of Statistics, 6, 65–70.

5. Hemming, K., Taljaard, M. & Forbes, A. Analysis of cluster randomised stepped wedge trials with repeated cross-sectional samples. Trials 2017;18:101 10.1186/s13063-017-1833-7

6. Cro S, Morris TP, Kenward MG, Carpenter JR. Sensitivity analysis for clinical trials with missing continuous outcome data using controlled multiple imputation: a practical guide. Stat Med 2020;39(21):2815–42. doi:10.1002/sim.8569

